# VALIDATION OF PROGRESS, A SIMPLE MACHINE-LEARNING DERIVED RISK STRATIFICATION SCORE FOR CASTRATION-RESISTANT PROSTATE CANCER

**DOI:** 10.64898/2026.02.24.26346978

**Authors:** L Castro Labrador, R Zamora, S Szyldergemajn, P Gómez del Campo, J Castillo-Izquierdo, JA De All, JM Domínguez, CM Galmarini

**Affiliations:** Topazium Artificial Intelligence, Madrid, Spain; Medicus, Buenos Aires, Argentina; DLPHI Oncology Global Consultants, Vaud, Switzerland

## Abstract

**Purpose:** Castration-resistant prostate cancer (CRPC) is characterized by marked clinical heterogeneity and poor long-term survival, underscoring the need for tools that can rapidly and reliably individualize patient risk. While several prognostic models exist, their complexity has limited routine clinical use. Here, we developed and validated PROGRESS (PROstate cancer Global Risk Evaluation and Stratification Score), a simplified prognostic score, derived through machine learning-guided feature selection, to enhance risk stratification and support individualized, risk-informed clinical decision-making.

**Methods:** PROGRESS was developed using baseline data from 2,035 metastatic CRPC patients enrolled in four different phase III trials. An unsupervised machine-learning approach was applied to identify latent patient subgroups with distinct survival outcomes irrespectively of allocated treatment arm, followed by classical multivariable modelling to derive a simple and straight-forward prognostic score based on routinely available objective laboratory variables. External validation was performed in three independent datasets comprising metastatic CRPC patients treated across different therapeutic settings (n=1,239) and non-metastatic CRPC patients managed with standard care (n=660). Overall survival was assessed using Kaplan-Meier and Cox regression analyses.

**Results:** Unsupervised modelling identified two patient risk subpopulations with significantly different overall survival rates (median 27.4 vs 17.7 months; hazard ratio [HR] 2.20, 95% CI 1.91–2.54; p<.00001). Feature contribution analysis yielded three independent predictors -PSA, ALP, and AST-used to build PROGRESS. In the training cohort, PROGRESS demonstrated strong discrimination (AUC 0.89). Using a prespecified cut-off, patients classified as increased risk had significantly shorter survival than low-risk patients (median 18.3 vs 25.6 months; HR 1.72, 95% CI 1.50–1.97; p<.0001). PROGRESS prognostic performance was consistent across all validation cohorts, including metastatic and non-metastatic disease, with HRs ranging from 1.74 to 3.46 (all p<.0001).

**Conclusions:** By integrating machine-learning-based pattern discovery with classical statistical modelling, PROGRESS provides a simple, objective, and clinically accessible approach for individual risk stratification in CRPC. Its reliance on three inexpensive, routinely measured laboratory parameters would facilitate practical implementation in clinical settings, enhancing visibility of underlying disease aggressiveness for individual clinical decision-making. PROGRESS could represent a pragmatic first step toward improving patient selection for clinical trials while identifying regulatory meaningful endpoints achievable in a sizeable patient population; further validation in prospective clinical studies and real-world datasets would allow to confirm its clinical utility and generalizability. PROGRESS can be freely accessed for research use only at the following link: https://dev.ai.topazium.com.

## INTRODUCTION

Castration-resistant prostate cancer (CRPC) typically emerges after an initial response to androgen deprivation therapy, during which tumours acquire adaptive resistance mechanisms that sustain malignant proliferation and survival under low-androgen conditions^1–3^. Treatment of metastatic CRPC (mCRPC) is multimodal and usually tailored according to prior systemic therapies, extent of disease, molecular characteristics, and the patient’s age and general health condition including frequently a non-negligible comorbidity profile^4–6^. It integrates continued androgen-deprivation therapy with last generation androgen receptor pathway inhibitors, taxane-based chemotherapy, immune chekpoint and PARP inhibitors in specific subtypes, as well as precision therapies, such as PSMA- or bone-targeted radioligand therapies when suitable^7–13^. These new therapies led a gradual but sustained improvement in median overall survival (OS) in randomized trials from roughly 16 months with mitoxantrone, currently limited to palliative use in heavily pretreated patients, to approximately 35 months with combination therapies^14–16^. Despite these advances, many studies fail to demonstrate a clear objective benefit over the evolving standard of care, largely because patients’ clinical heterogeneity only becomes apparent after long and costly follow-up. In this context, there is a critical unmet need to rapidly and reliably identify patients with particularly poor prognosis, both to improve patient selection for clinical trials and to accurately determine the early need for more intensive or aggressive therapeutic strategies only in those patients most likely to benefit.

To date, several prognostic models based on routinely available clinical variables (e.g., visceral involvement, performance status, PSA level, etc.) have been developed to estimate patient survival and stratify risk of death^17–19^. For instance, the Halabi model classifies patients into low-, intermediate-, and poor-risk groups and provides robust estimates of 1- and 2-year survival^18^. However, despite their demonstrated prognostic performance and utility in clinical trials, the relative complexity of these models and their limited integration into routine clinical workflows have constrained their widespread adoption for rapid, individualized patient-level risk assessment. There remains an unmet need for tools that can efficiently assess both the risk of death and the likelihood of relatively uneventful long survival versus an aggressive disease course early on, using simpler and more easily applicable prognostic instruments.

To address this need, we developed and evaluated a simplified PROstate cancer Global Risk Evaluation and Stratification Score (PROGRESS), an accessible, sensitive predictive tool that utilizes three common laboratory metrics to identify CRPC patients at high risk of death. We first applied machine learning (ML) techniques to uncover patterns and features associated with patient clinical outcome, which were then used as input variables to build the predictive score. The score was then validated using a large cohort of patients from five distinct clinical trials involving CRPC patients at non-metastatic (M0) or metastatic (M1) stages. Our findings suggest that PROGRESS accurately identifies, those patients who are likely to have a high-risk disease with likely limited survival.

## METHODS

### Patient Population and datasets

The patient cohort comprised individuals with prostate cancer enrolled in the following clinical trials: NCT00273338, NCT00988208, NCT00617669, NCT00519285, NCT00554229, NCT00676650, TAX327, NCT00417079 and NCT00626548 (www.clinicaltrials.gov). Detailed descriptions of these studies are available in Supplementary Table 1. All datasets were sourced from www.projectdatasphere.org. The training set (TRS) included 2,035 patients from NCT00273338 (N=463), NCT00988208 (N=511), NCT00617669 (N=457), and NCT00519285 (N=604)^20–23^. These patients all had mCRPC and were treated with docetaxel combined with prednisone.

As validation sets (VS), we used five independent clinical trials joined in three different datasets. VS1 comprised mCRPC post-first-line chemotherapy-treated patients from the placebo arm of the NCT00554229 (N=256) and the NCT00676650 (N=275) clinical trials^24,25^. These patients received standard-of-care treatment, including steroids (prednisone or prednisolone), bisphosphonates, pain management therapies, and surgical or continuous medical castration. VS2 included patients treated with mitoxantrone as first- or second-line therapy from the control arms of the TAX327 (N=335) and the NCT00417079 (N=373) clinical trials^26,27^. Finally, VS3 (N=660) comprised non-metastatic (M0) CRPC patients from the placebo arm of NCT00626548, characterized by rising serum PSA levels despite medical or surgical castration and managed with standard care, which could include symptomatic treatment for urinary obstruction, surgical or medical castration, prednisolone, ketoconazole, or megestrol acetate^28^.

### Input Data

In the training set, 161 variables were available (listed in Supplementary Table 2) for all patients, collected during the screening phase of trial enrolment. For categorical variables, missing values were treated as negatives. For quantitative variables, missing values were imputed using our proprietary Signal Healer^TM^ tool, a graph neural network for robust signal reconstruction, freely accessible at https://dev.ai.topazium.com. For individuals in the validation sets (VS), data included PSA, ALP, AST values and OS data. On these datasets, no missing values were accepted.

### ML Model and predictive Score Development

Patient data were integrated into feature vectors to generate synthetic representations (SRs) that subsequently were analysed using UMAP (umap-learn v0.5.3) for dimensionality reduction and clustered via DBSCAN (scikit-learn v1.2.2). Resultant subpopulations were correlated with OS, and statistically significant variables (p<.05) were identified through feature contribution analysis. These were used to create PROGRESS as previously described elsewhere^29,30^. For this purpose, baseline values of statistically significant variables in the feature contribution analysis were incorporated into the logistic regression model, employing a backward stepwise approach to derive the final predictors. The independent predictive factors that retained statistical significance in the final logistic regression analysis were utilized to compute the predictive score derived from hazard ratios (HR). Model performance was assessed using the area under the receiver operating characteristic (ROC) curve and the Hosmer-Lemeshow test. The optimal cut-off value for the score was determined using Youden’s index, complemented by an evaluation of the accuracy at that threshold to ensure robust classification.

### Statistical Analysis

Statistical tests included chi-square for categorical and Mann-Whitney U for continuous variables. Kaplan-Meier analysis, performed with the survival package (v3.5-7) in R (v4.3.2), was used for OS estimation that was calculated from the date of randomization to the date of death from any cause, respectively, or to the last follow-up examination (censored event). Differences with p<.05 were deemed statistically significant.

### PROGRESS website tool

PROGRESS can be freely accessed for research use only at the following link: https://dev.ai.topazium.com.

## RESULTS

### Development of an Unsupervised ML Model

Using SRs from 2,035 training-set patients (baseline characteristics in Supplementary Table 3 and Supplementary Figure 1), an unsupervised machine-learning model stratified the cohort into two subgroups, SPA (N=828; 40.6%) and SPB (N=1,207; 59.4%) (Figure 1). Median OS was 27.4 months (CI95%: 25.8-30.4) in SPA and 17.7 months (CI95%: 16.8-18.5) in SPB (Figure 2A). Consistent with these estimates, survival analysis demonstrated a more than two-fold higher risk of death for SPB versus SPA (HR: 2.20; CI95%: 1.91-2.54; p<.00001) patients.

**Figure 1.**
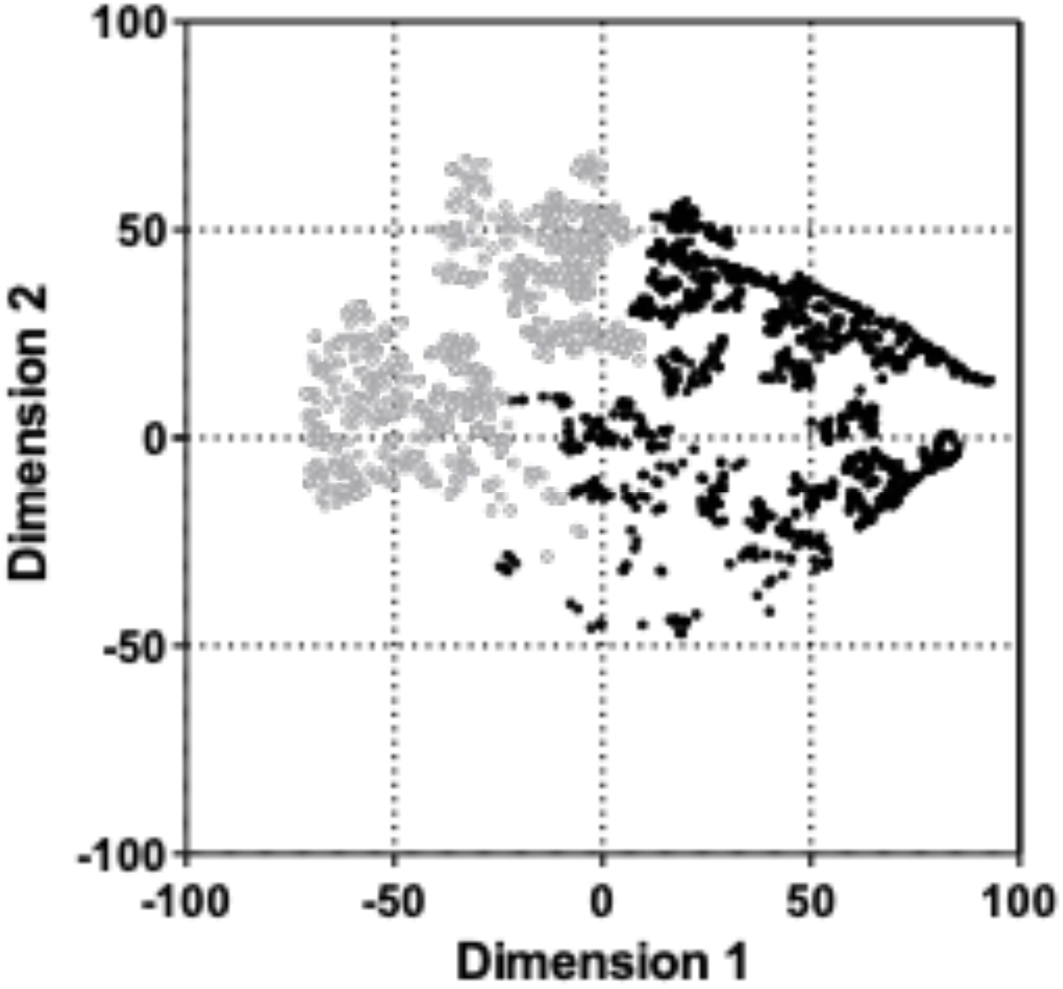
Embedding space and clustering of patients belonging to the training dataset by a machine learning framework. Each dot in the graph corresponds to the synthetic representation (SR) of each patient of the dataset. The SR was obtained through the integration of 161 clinical and analytical features collected, respectively, during the screening phase and the first month of inclusion on the trial. Two major clusters were identified: subpopulation A (SPA; n=828; grey, open dots) and subpopulation B (SPB; n=1,207; black, solid dots).

**Figure 2.**
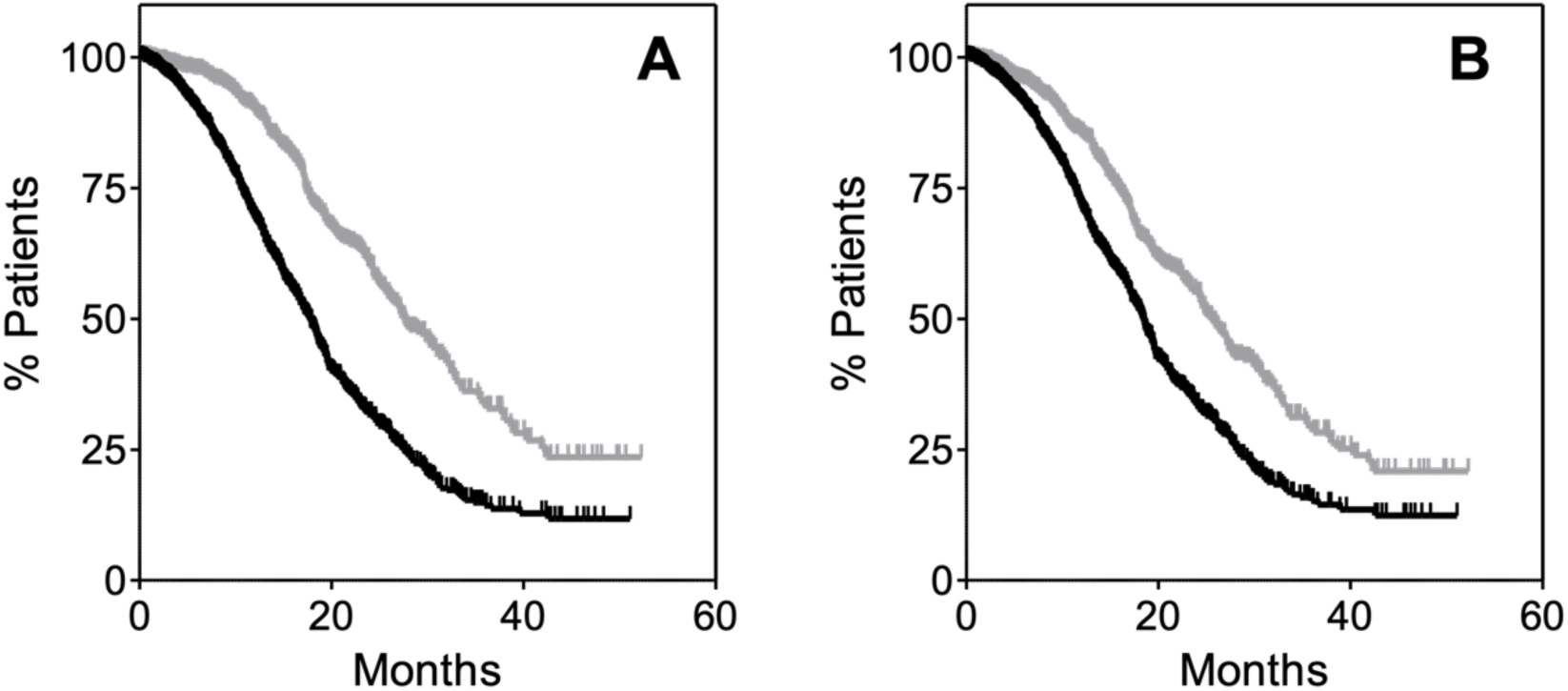
Kaplan-Meier estimates of overall survival (OS) in the training set based on (A) the unsupervised machine learning model and (B) PROGRESS. In panel (A), SPA patients are represented in grey and SPB patients in black. In panel (B), patients with PROGRESS <34 are shown in grey and PROGRESS ≥34 are shown in black.

Feature contribution analysis identified 43 key variables that differentiated patients in SPA from those in SPB. As shown in Supplementary Table 4, some clinical and laboratory features were distinct between the two groups. SPB comprised a larger proportion of patients than SPA (59.1% vs 40.9%). Relative to SPA, patients in SPB had slightly lower body surface area and BMI, but higher heart rate and both systolic and diastolic blood pressure (all p<.01). Differences in some serum markers were pronounced: PSA, LDH and ALP levels were substantially higher in SPB, as were AST values, whereas albumin and haemoglobin were lower (all p<.001). Prior orchidectomy was more common in SPB, whereas TURP and leuprolide use were more frequent in SPA. Bone-related signs and symptoms, including bone pain, were more frequent in SPB (p<.01), which was consistent with higher use of bone-targeted agents and analgesics such as bisphosphonates (including zoledronic acid), total opioids, natural opium alkaloids, other opioids, and opioid-analgesic combinations (all p≤.005). In contrast, biguanides, lipid-modifying agents such as HMG-CoA reductase inhibitors (particularly simvastatin), and ACE inhibitors were more commonly used in SPA (p<.01).

### Generation of the PROGRESS Score

The PROGRESS score was developed to identify CRPC patients at higher risk of death, using a smaller set of clinically accessible variables than those highlighted by the ML framework. Baseline values of the 43 variables differentiating the SPA and SPB subpopulations were entered into a multivariable logistic regression model, which identified PSA, ALP, and AST as independent predictors of outcome (Table 1). The PROGRESS score was derived from the HRs of these predictors, rounded to the nearest integer or 0.5 as appropriate, yielding a score ranging from 0 to 76.5 points, with 42.5 points assigned for PSA >66 ng/mL, 30.5 for ALP >127 IU/L, and 3.5 points for AST >32 IU/L. In the training set, PROGRESS yielded an AUC of 0.89 (95% CI 0.88-0.91) for predicting survival (Supplementary Figure 2).

**Table 1.** Proposed PROGRESS scoring system and clinical interpretation.

| Variable | $\beta$ | HR* (CI95%) | P value | Score points |
| --- | --- | --- | --- | --- |
| PSA >66 ng/ml | 3.75 | 42.7 (29.4-61.9) | <.001 | 42.5 |
| ALP >127 IU/L | 3.41 | 30.4 (21.1-43.9) | <.001 | 30.5 |
| AST >32 IU/L | 1.28 | 3.6 (2.48-5.29) | <.001 | 3.5 |
| Range of score |  |  |  | 0-76.5 |
| Total Score |  |  |  | Interpretation |
| $\geq 34$ | | | | Increased-risk of death |
| <34 |  |  |  | Low-risk of death |

The analysis was performed with all variables as described in Materials and Methods.
Variables not shown here yielded non-significant P-values ( $>.05$ )
\* HR: hazard ratio; PSA: prostate specific antigen; ALP: alkaline phosphatase; AST: aspartate transaminase

To dichotomize patients into increased- and low-risk patients, a threshold of 34 points was selected based on a Youden’s index of 0.62. Patients scoring ≥34 were classified as being at increased-risk of death, whereas those with a score <34 were classified as being low risk. This cut-off was subsequently applied to the training cohort for internal validation. For this analysis, we included only patients with complete, non-imputed values for all three PROGRESS variables resulting in 1,979 individuals, 97% of the original cohort. Descriptive statistics for PSA, ALP, and AST in the TRS are provided in Supplementary Table 5. Among them, 1,112 (56.1%) had a PROGRESS score ≥34 and 867 (43.8%) had a score <34. Survival analyses were then performed. As illustrated in Figure 2B, patients with PROGRESS ≥34 had a significantly higher risk of death (HR 1.72; CI95%: 1.50-1.97; p<.0001), with a median OS of 18.3 months (CI95%: 17.3-19.0), compared with a median OS of 25.6 months (CI95%: 23.9-27.2) in those with PROGRESS <34. These results confirmed that PROGRESS discriminated patients at increased risk of mortality.

### Validation of PROGRESS in three different validation sets of CRPC patients

To validate the PROGRESS results, we analysed three independent datasets of patients with CRPC, comprising two M1 cohorts and one M0 cohort. Descriptive statistics for PSA, ALP, and AST, together with OS analyses for the full population of each validation dataset, are summarized in Supplementary Table 5 and Supplementary Figure 3. The first validation dataset (VS1) included 531 mCRPC patients treated with standard of care after first-line therapy (see Materials and Methods). Among these patients, 314 (59.1%) had a PROGRESS score ≥34, whereas 217 (40.9%) had a PROGRESS score <34. Survival analysis showed that patients with PROGRESS ≥34 experienced a significantly shorter OS compared with those with PROGRESS <34 (HR: 2.61; 95% CI: 2.03-3.35; p<.0001) (Figure 3A). Median OS was 11.9 months (95% CI: 10.6-14.1) in the increased-risk group and was not reached in the low-risk group (95% CI: 19.9-not reached) (Table 2).

**Figure 3.**
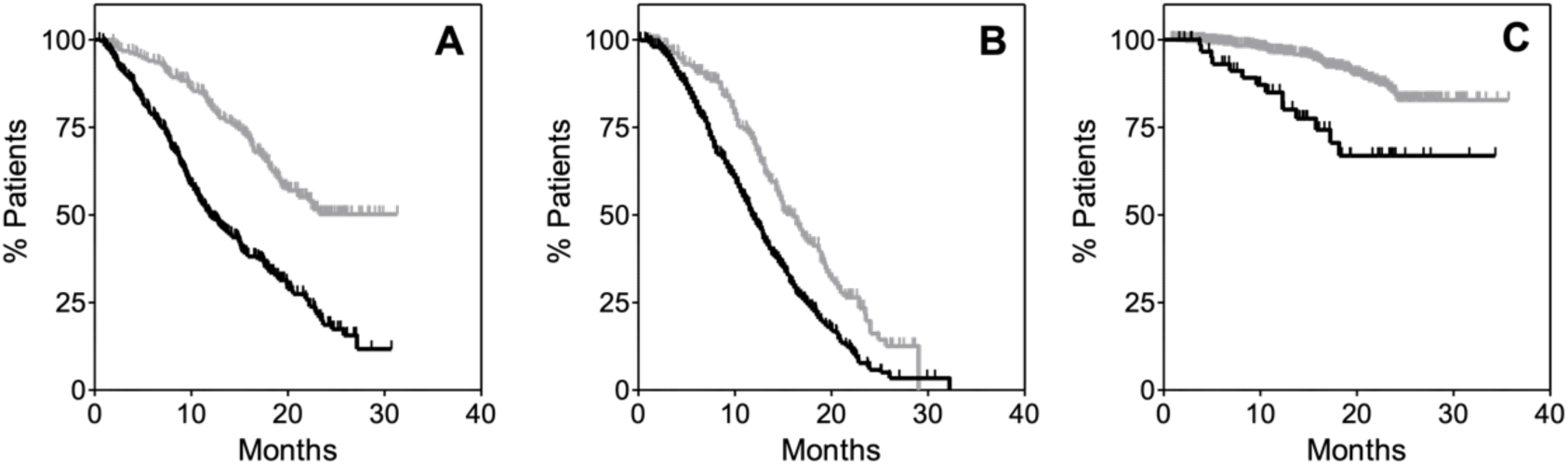
Kaplan-Meier estimates of overall survival (OS) in the validation sets stratified by PROGRESS. Panel (A) shows VS1, panel (B) VS2, and panel (C) VS3. In all panels, patients with PROGRESS ≥34 are shown in black, whereas patients with PROGRESS <34 are represented in grey.

**Table 2.** Summary of survival results for the validation datasets.

|  | N (%) | Median* (CI95%) | HR (CI95%) | P-value |
| --- | --- | --- | --- | --- |
| VS1 |  |  |  |  |
| ≥34 | 314 (59.1) | 11.9 (10.6-14.1) | 2.61 (2.03-3.35) | <.0001 |
| <34 | 217 (40.9) | NR** (19.9-NR) |  |  |
| VS2 |  |  |  |  |
| ≥34 | 492 (69.5) | 11.7 (11.1-12.9) | 1.74 (1.41-2.15) | <.0001 |
| <34 | 216 (30.5) | 15.9 (14.3-17.5) |  |  |
| VS3 |  |  |  |  |
| ≥34 | 65 (9.9) | NR (NR) | 3.46 (1.89-6.33) | <.0001 |
| <34 | 595 (90.1) | NR (NR) |  |  |

\* OS expressed in months
\*\* NR: not reached

We next evaluated the second validation dataset (VS2), which comprised 708 mCRPC patients treated with mitoxantrone plus prednisone as first- or second-line therapy (see Materials and Methods). Of these, 492 patients (69.5%) had PROGRESS ≥34 and 216 (30.5%) had PROGRESS <34. Consistent with the VS1 results, patients with PROGRESS ≥34 had significantly shorter OS than those with PROGRESS <34 (HR: 1.74; 95% CI: 1.41-2.15; p<.0001) (Figure 3B). Median OS was 11.7 months (95% CI: 11.1-12.9) in the increased-risk group and 15.9 months (95% CI: 14.3-17.5) in the low-risk group (Table 2). Finally, we assessed PROGRESS performance in a cohort of 660 patients with non-metastatic CRPC (M0 stage; see Materials and Methods). In this dataset, 65 patients (9.9%) had PROGRESS ≥34, while 595 (90.1%) had PROGRESS <34. Survival analysis again demonstrated a significantly shorter OS for patients with PROGRESS ≥34 compared with those with PROGRESS <34 (HR: 3.46; 95% CI: 1.89-6.33; p<.0001) (Figure 3C and Table 2).

## DISCUSSION

Herein, we report the results from the development and validation of PROGRESS, a simple and clinically applicable prognostic score designed to categorize survival risk in patients with CRPC. PROGRESS is based exclusively on baseline values of three routinely available, objective, and inexpensive laboratory parameters -PSA, ALP, and AST-which would make it straightforward to be implemented and suitable for use in everyday clinical practice, including ambulatorily and primary care by general practitioners (GPs). These variables were identified through the combined application of unsupervised machine-learning techniques and classical multivariable biostatistical modelling, allowing the initial high-dimensional ML output to be translated into a clinically interpretable score. Despite its simplicity, the three-predictor PROGRESS model demonstrated strong discriminative performance in the training cohort and showed consistent prognostic accuracy across multiple independent validation datasets, spanning both metastatic and non-metastatic disease stages. Importantly, this accuracy was also observed in patients treated across different lines of therapy as well as individuals managed only with symptomatic or palliative care or with no treatment at all, underscoring its broad applicability and clinical utility. While PROGRESS does not capture the full spectrum of molecular, or molecular-related biological factors involved in predicting responses to specific therapies, it unveils the potential for purely prognostic features and its simplicity support its use as a practical tool for rapid risk assessment and patient stratification in settings where ease of use and timely decision-making are essential. In this context, PROGRESS aligns with a precision medicine approach aimed at improving patient stratification and optimizing clinical management, while avoiding unnecessary interventions in patients with more favourable prognoses.

As described, PROGRESS was developed using a sequential and integrative strategy that combined ML techniques with classical statistical modelling, with the primary goal of improving risk visibility at the individual patient level. Specifically, an unsupervised ML approach was first applied to identify latent subpopulations of CRPC patients characterized by distinct clinical and biological profiles associated with differential survival outcomes. This data-driven step leveraged the ability of advanced analytics to detect complex, multivariate patterns of individual heterogeneity relevant to the disease natural history, in a rather controlled environment of clinical trials, without relying on or being biased by predefined clinical assumptions. Subsequently, multivariable regression analysis was used to distil these high-dimensional ML findings into a transparent and clinically interpretable prognostic score based on a limited set of routinely available laboratory variables. This approach allowed us to leverage the pattern-recognition power of a “black-box” system, while ultimately providing a “clear-box” tool that clinicians are familiar with and can readily apply in routine practice. By translating unsupervised ML outputs into a simple scoring system, PROGRESS reduces the uncertainty often associated with complex computational models and bridges the gap between advanced analytics and clinical usability. Noteworthy, the unsupervised framework revealed clinically meaningful differences between patient subgroups, including variations in laboratory parameters both above and within normal ranges, underscoring the model’s ability to capture intrinsic prognostic features that are not readily discernible through conventional clinical judgement alone. We have previously used this strategy successfully. In stage IV squamous NSCLC, high-dimensional clinical-laboratory fingerprints uncovered subpopulations with distinct outcomes and toxicity profiles to first-line treatment with gemcitabine and cisplatin, and in colorectal cancer, the PROCC score effectively detected KRAS wild-type metastatic patients most likely to benefit from EGFR-directed therapies^29,31^. These prior results support the feasibility and impact of leveraging multidimensional clinical-laboratory signatures combined with a classical scoring system, reinforcing the rationale for applying the same approach in this study.

Several prognostic models have been developed to estimate survival and stratify risk in advanced prostate cancer, among which the Halabi model remains one of the most extensively validated and widely used, particularly in clinical trials^18^. In this context, PROGRESS was deliberately developed as a simplified prognostic tool based on three routinely available and objective laboratory parameters (PSA, ALP, and AST), with the goal of enabling timely and reproducible identification of patients’ risk without introducing interobserver differences or clinical biases. Importantly, PROGRESS is straightforward to use and is currently accessible through a freely available online platform, facilitating its application in both research and clinical settings. In parallel, the development of a mobile application is underway to further support prospective clinical validation and real-world implementation. PROGRESS is not intended to replace established prognostic models, but rather to complement them by offering a pragmatic option for early risk assessment and for guiding personalized medical treatments in settings that demands rapid decisions.

PSA, ALP, and AST emerged as the key variables underpinning the PROGRESS score. These markers are not novel individually; rather, their combined and weighted integration into a single score appears to enhance prognostic discrimination beyond that achieved by any individual parameter alone. While the biological relevance of each variable is well supported by existing literature, the precise mechanisms by which their interaction delineates distinguishable survival projections remain to be fully understood. There is accumulating evidence that PSA kinetics have prognostic value in patients with CRPC^32–37^. Increased ALP levels are a surrogate marker for skeletal involvement and bone turnover, a central feature of advanced CRPC^38–40^. AST, although less commonly emphasized in prognostic models, may capture aspects of early hepatic involvement, inflammation, and its role in cancer metabolic pathways including the Warburg effect or glutamine production^41,42^. This underscores the need for rationally guided hypothesis generating translational studies to elucidate the underlying biological pathways linking these laboratory abnormalities to disease clinical behaviour and improving outcomes in CRPC clinical studies.

This study has several limitations that warrant acknowledgment. First, PROGRESS was developed using retrospective analyses of clinical trial datasets, which inherently reflect a relatively selected patient environment and may limit generalizability to broader real-world populations. For example, patients with poor performance status (e.g., ECOG ≥2) were underrepresented, and laboratory values showed a narrower distribution, as extreme outliers would have been excluded under typical trial eligibility criteria. Nevertheless, the included datasets exhibited sufficient clinical heterogeneity in key prognostic variables, including PSA levels and metastatic status (M0 vs. M1), enabling robust model development and validation. Second, most of the source trials were conducted prior to the widespread adoption of more contemporary agents for CRPC, and therefore the observed survival patterns may not fully reflect clinical outcomes under newer treatment paradigms. Consequently, they may underestimate the improvements in standard supportive care over the past decade. Third, the score was derived using a complete-case approach without imputation, resulting in the exclusion of patients with missing baseline PSA, ALP, or AST values in certain analyses; while this may introduce some bias, the impact appeared limited, as consistent performance was observed across independent validation cohorts. Finally, PROGRESS is a purely prognostic model and does not incorporate treatment-predictive models of specific subsets, molecular alterations, or longitudinal biomarker dynamics, which could further refine risk stratification in some specific settings (e.g., MSI-H or HRD features). Despite these limitations, PROGRESS represents a clinically accessible pure prognostic tool, developed using complementary machine-learning and classical statistical approaches and demonstrating stable performance across multiple clinical trial datasets.

In conclusion, PROGRESS is a simple, robust, and clinically interpretable prognostic score that enables effective mortality risk stratification of patients with CRPC using only three inexpensive, routinely available laboratory parameters. Developed through an integrative approach combining unsupervised machine learning with classical statistical modelling, PROGRESS consistently identified patient subgroups with markedly different survival outcomes across multiple independent cohorts and disease stages. Its ease of implementation and strong reproducibility support its potential utility as a pragmatic tool to complement existing prognostic frameworks in clinical practice. Future prospective studies and real-world validations will be essential to further define its role in guiding clinical decision-making and optimizing patient management in CRPC.

## Data Availability

All data produced are available online at http://www.projectdatasphere.com/

## ACKNOWLEDGEMENTS

We are grateful to http://www.projectdatasphere.com/ that allow us to produce this research through access to the high-quality datasets used in the study.

## COMPETING INTERESTS

LCL, PGC, JCI and JMD are employees of Topazium. CMG is a shareholder and employee of Topazium. SS is a shareholder and employee of DLPHI Oncology Global Consultants. JADA is a shareholder and employee of Medicus. RZ is an employee of Medicus.

## SUPLEMENTARY TABLES

**Supplementary Table 1.** List of clinical studies used as datasets for development and validation of PROGRESS.

| NCT term | Study type | Arm | Description |
| --- | --- | --- | --- |
| NCT00273338 | Phase III | Docetaxel | Calcitriol in combination with docetaxel in androgen-independent prostate cancer |
| NCT00988208 | Phase III | Docetaxel | Lenalidomide in combination with docetaxel and prednisone for patients with castrate-resistant prostate cancer |
| NCT00617669 | Phase III | Docetaxel | Zibotentan and docetaxel in metastatic hormone resistant prostate cancer |
| NCT00519285 | Phase III | Docetaxel | Aflibercept in combination with docetaxel in metastatic androgen independent prostate cancer |
| NCT00554229 | Phase III | Placebo | Zibotentan in hormone resistant prostate cancer with bone metastases |
| NCT00676650 | Phase III | Placebo | Sunitinib plus prednisone vs. prednisone in patients with progressive metastatic castration-resistant prostate cancer after failure of a docetaxel-based chemotherapy |
| TAX327 | Phase III | Mitoxantrone | Docetaxel administered either weekly or every three weeks vs. mitoxantrone for metastatic hormone refractory prostate cancer |
| NCT00417079 | Phase III | Mitoxantrone | Cabazitaxel vs. mitoxantrone in combination with prednisone for the treatment of hormone refractory metastatic prostate cancer previously treated with a docetaxel-containing regimen |
| NCT00626548 | Phase III | Placebo | Zibotentan in non-metastatic hormone resistant prostate cancer |

**Supplementary Table 2.** List of variables used as input data on the training set.

| Variable | Units |
| --- | --- |
| Age | years at screening |
| Weight | Kg |
| Height | cm |
| BSA* | m<sup>2</sup> |
| BMI | Kg/m<sup>2</sup> |
| Race | (value; 0-5) |
| ECOG performance status | (value; 0-4) |
| Overall survival | months |
| Death | Y/N (1,0) |
| Pulse (bpm) | (value; bpm) |
| DBP | (value; mmHg) |
| SBP | (value; mmHg) |
| Previous surgery | Y/N (1,0) |
| Abscess drainage | Y/N (1,0) |
| Aortic valve replacement | Y/N (1,0) |
| Appendicectomy | Y/N (1,0) |
| Arterial stent insertion | Y/N (1,0) |
| Arteriovenous fistula operation | Y/N (1,0) |
| Aspiration biopsy | Y/N (1,0) |
| Benign tumour excision | Y/N (1,0) |
| Bilateral orchidectomy | Y/N (1,0) |
| Biopsy liver | Y/N (1,0) |
| Biopsy lymph gland | Y/N (1,0) |
| Biopsy prostate | Y/N (1,0) |
| Biopsy stomach | Y/N (1,0) |
| Bladder neoplasm surgery | Y/N (1,0) |
| Bladder polypectomy | Y/N (1,0) |
| Brachytherapy to prostate | Y/N (1,0) |
| Cardiac pacemaker insertion | Y/N (1,0) |
| Cataract operation | Y/N (1,0) |
| Cholecystectomy | Y/N (1,0) |
| Colectomy | Y/N (1,0) |
| Colectomy total | Y/N (1,0) |
| Colonoscopy | Y/N (1,0) |
| Colostomy | Y/N (1,0) |
| Coronary arterial stent insertion | Y/N (1,0) |
| Coronary artery bypass | Y/N (1,0) |
| Cryotherapy | Y/N (1,0) |
| Cystoscopy | Y/N (1,0) |
| Cystostomy | Y/N (1,0) |
| Explorative laparotomy | Y/N (1,0) |
| Foot fracture | Y/N (1,0) |
| Gastrectomy | Y/N (1,0) |
| Gastric operation | Y/N (1,0) |
| Gastric ulcer surgery | Y/N (1,0) |
| Haemorrhoid operation | Y/N (1,0) |
| Hernia repair | Y/N (1,0) |
| Hip arthroplasty | Y/N (1,0) |
| Hydrocele operation | Y/N (1,0) |
| Implantable defibrillator insertion | Y/N (1,0) |
| Inguinal hernia repair | Y/N (1,0) |
| Intervertebral disc operation | Y/N (1,0) |
| Knee arthroplasty | Y/N (1,0) |
| Lenticular operation | Y/N (1,0) |
| Lipoma excision | Y/N (1,0) |
| Lymphadenectomy | Y/N (1,0) |
| Malignant tumour excision | Y/N (1,0) |
| Mastoidectomy | Y/N (1,0) |
| Nephrectomy | Y/N (1,0) |
| Nephrostomy | Y/N (1,0) |
| Oesophageal operation | Y/N (1,0) |
| Orchidectomy | Y/N (1,0) |
| Orthopedic procedure | Y/N (1,0) |
| Osteotomy | Y/N (1,0) |
| Prostatectomy | Y/N (1,0) |
| Radical prostatectomy | Y/N (1,0) |
| Resection of rectum | Y/N (1,0) |
| Retro-pubic prostatectomy | Y/N (1,0) |
| Rhinoplasty | Y/N (1,0) |
| Shoulder operation | Y/N (1,0) |
| Sigmoidectomy | Y/N (1,0) |
| Sinus operation | Y/N (1,0) |
| Skin neoplasm excision | Y/N (1,0) |
| Spinal cord operation | Y/N (1,0) |
| Spinal laminectomy | Y/N (1,0) |
| Suprapubic prostatectomy | Y/N (1,0) |
| Surgery | Y/N (1,0) |
| Thoracotomy | Y/N (1,0) |
| Tonsillectomy | Y/N (1,0) |
| Total adrenalectomy | Y/N (1,0) |
| Transurethral bladder resection | Y/N (1,0) |
| Transurethral prostatectomy | Y/N (1,0) |
| Umbilical hernia repair | Y/N (1,0) |
| Urethral stent insertion | Y/N (1,0) |
| Urinary cystectomy | Y/N (1,0) |
| Varicose vein operation | Y/N (1,0) |
| Vascular graft | Y/N (1,0) |
| Vascular stent insertion | Y/N (1,0) |
| Vertebroplasty | Y/N (1,0) |
| Vocal cordectomy | Y/N (1,0) |
| Previous cancer therapy (any) | Y/N (1,0) |
| 5-α-reductase inhibitors | Y/N (1,0) |
| Antiandrogens | Y/N (1,0) |
| Chemotherapy | Y/N (1,0) |
| CYP17 inhibitors | Y/N (1,0) |
| Estrogen analogues | Y/N (1,0) |
| Estrogens | Y/N (1,0) |
| Experimental | Y/N (1,0) |
| Glucocorticoids | Y/N (1,0) |
| GnRH agonists | Y/N (1,0) |
| Imidazole derivatives | Y/N (1,0) |
| mTOR inhibitors | Y/N (1,0) |
| Progestogens | Y/N (1,0) |
| Radioisotope | Y/N (1,0) |
| Somatostatin analogues | Y/N (1,0) |
| Tyrosine-kinase inhibitors | Y/N (1,0) |
| Prior hormonotherapy | Y/N (1,0) |
| Prior antiandrogens | Y/N (1,0) |
| Abiraterone | Y/N (1,0) |
| Bicalutamide | Y/N (1,0) |
| Cyproterone | Y/N (1,0) |
| Diethylstilbestrol | Y/N (1,0) |
| Dienestrol | Y/N (1,0) |
| Dutasteride | Y/N (1,0) |
| Enzalutamide | Y/N (1,0) |
| Estradiol | Y/N (1,0) |
| Finasteride | Y/N (1,0) |
| Fluconazole | Y/N (1,0) |
| Flutamide | Y/N (1,0) |
| Genistein | Y/N (1,0) |
| Hexestrol | Y/N (1,0) |
| Ketoconazole | Y/N (1,0) |
| Megestrol | Y/N (1,0) |
| Nilutamide | Y/N (1,0) |
| Polyestradiol | Y/N (1,0) |
| Tamoxifen | Y/N (1,0) |
| Previous GnRH therapy | Y/N (1,0) |
| Abarelix | Y/N (1,0) |
| Buserelin | Y/N (1,0) |
| Degarelix | Y/N (1,0) |
| Gonadorelin | Y/N (1,0) |
| Goserelin | Y/N (1,0) |
| Histrelin | Y/N (1,0) |
| Leuprolide | Y/N (1,0) |
| Previous radiotherapy | Y/N (1,0) |
| Previous chemotherapy | Y/N (1,0) |
| Cabazitaxel | Y/N (1,0) |
| Cabozantinib | Y/N (1,0) |
| Cyclophosphamide | Y/N (1,0) |
| Dasatinib | Y/N (1,0) |
| Docetaxel | Y/N (1,0) |
| Estramustine | Y/N (1,0) |
| Etoposide | Y/N (1,0) |
| Mitoxantrone | Y/N (1,0) |
| Raloxifene | Y/N (1,0) |
| Prior-Bone-modifying agents | Y/N (1,0) |
| Denosumab | Y/N (1,0) |
| Biphosphonates (any) | Y/N (1,0) |
| Zoledronic acid | Y/N (1,0) |
| Medical history | (ICD10 code) |
| Albumin | g/L |
| ALT | UI/L |
| ALP | UI/L |
| AST | UI/L |
| Creatinine | μmol/L |
| Hemoglobin | g/L |
| LDH | UI/L |
| PSA | ng/mL |
| Testosterone | ng/mL |
| Concomitant medication | ATC3 code |
| Opiates | ATC3 code |

BSA: body surface area; BMI: body mass index; DBP: diastolic blood pressure; SBP: systolic blood pressure; ALT: alanine transaminase; ALP: alkaline phosphatase; AST: aspartate aminotransferase; LDH: lactate dehydrogenase; PSA: prostate specific antigen; Y: yes; N: no.

**Supplementary Table 3.** Main clinical characteristics of patients on the training dataset (N=2,035).

| Variable | Mean | CI95% |
| --- | --- | --- |
| Age (years) | 69.1 | 68.6-69.7 |
| Weight (Kg) | 89.0 | 82.2-90.0 |
| Height (cm) | 1.742 | 1.739-1.746 |
| BSA* (m<sup>2</sup>) | 2.03 | 2.027-2.046 |
| BMI | 28.0 | 27.8-28.2 |
| Pulse (bpm) | 75.2 | 74.2-76.2 |
| DBP (mmHg) | 78.0 | 77.3-78.6 |
| SBP (mmHg) | 133.4 | 132.4-134.4 |
| Albumin (g/L) | 4.18 | 4.16-4.19 |
| ALP (UI/L) | 242.9 | 228.8-257.5 |
| ALT (UI/L) | 23.1 | 22.7-23.5 |
| AST (UI/L) | 26.7 | 26.3-27.1 |
| Creatinine (μmol/L) | 0.99 | 0.97-1.00 |
| LDH (UI/L) | 235.3 | 229.7-241.0 |
| Hemoglobin (g/L) | 12.64 | 12.59-12.69 |
| PSA (ng/mL) | 237.5 | 200.9-265.9 |
| Testosterone (ng/mL) | 0.61 | 0.51-0.71 |
|  | N | % |
| Clinical Study |  |  |
| NCT00273338 | 519 | 24.8 |
| NCT00988208 | 511 | 24.4 |
| NCT00617669 | 457 | 21.8 |
| NCT00519285 | 604 | 28.8 |
|  | N | Mean % |
| White race | 1,729 | 82.6 |
| ECOG 0/1 | 2,006 | 95.9 |

\* All patients had metastatic castration-resistant prostate cancer and were treated in the control arm of the study with docetaxel (75 mg/m<sup>2</sup> BSA) intravenously on day 1 of a 21-day cycle, combined with continuous prednisone 5 mg PO (BID).
\*\* BSA: body-surface area; BMI: body mass index; DBP: diastolic blood pressure; SBP: systolic blood pressure; PSA: prostate-specific antigen; LDH: lactate dehydrogenase; ALP: alkaline phosphatase; AST: aspartate transaminase.

**Supplementary Table 4.**
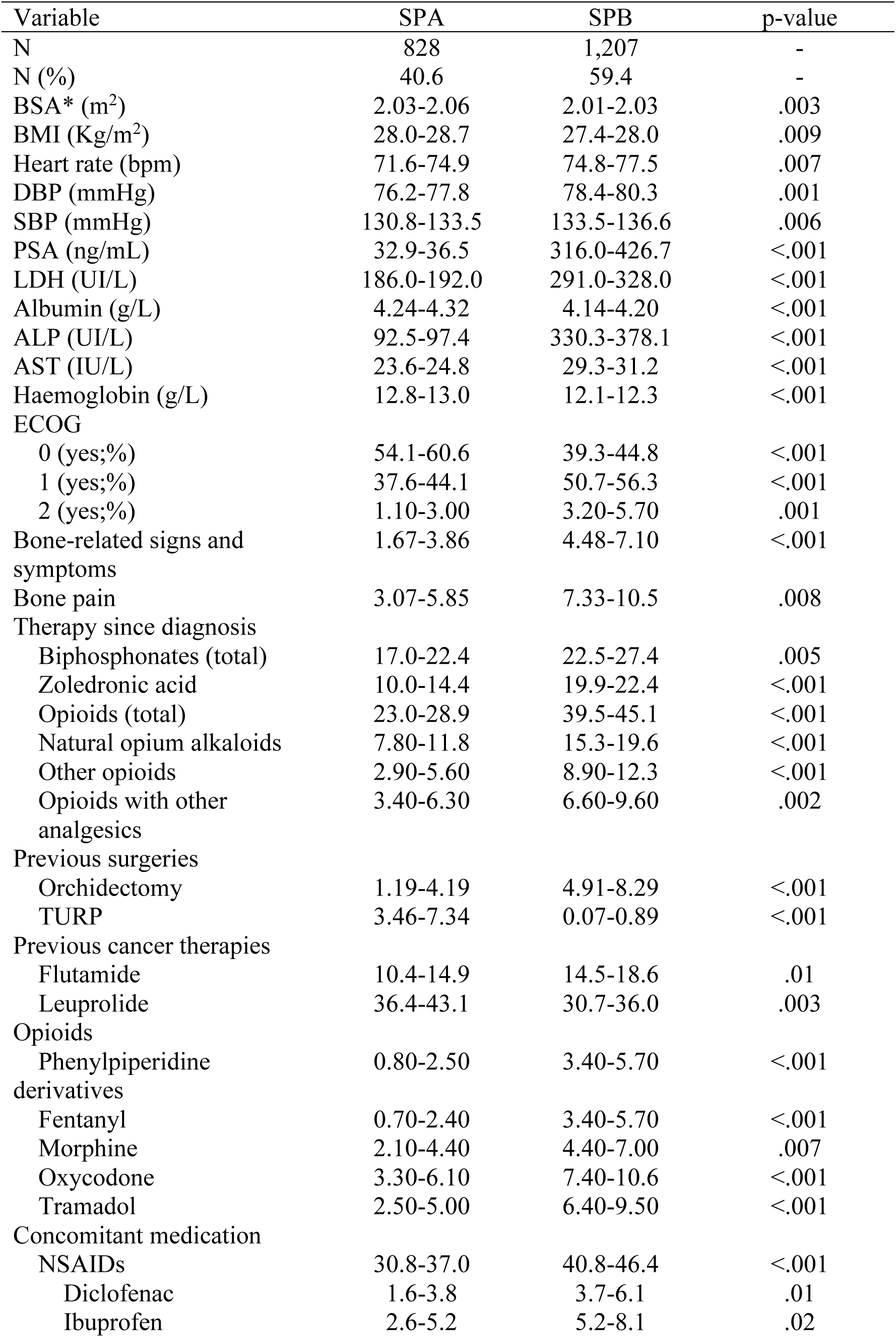
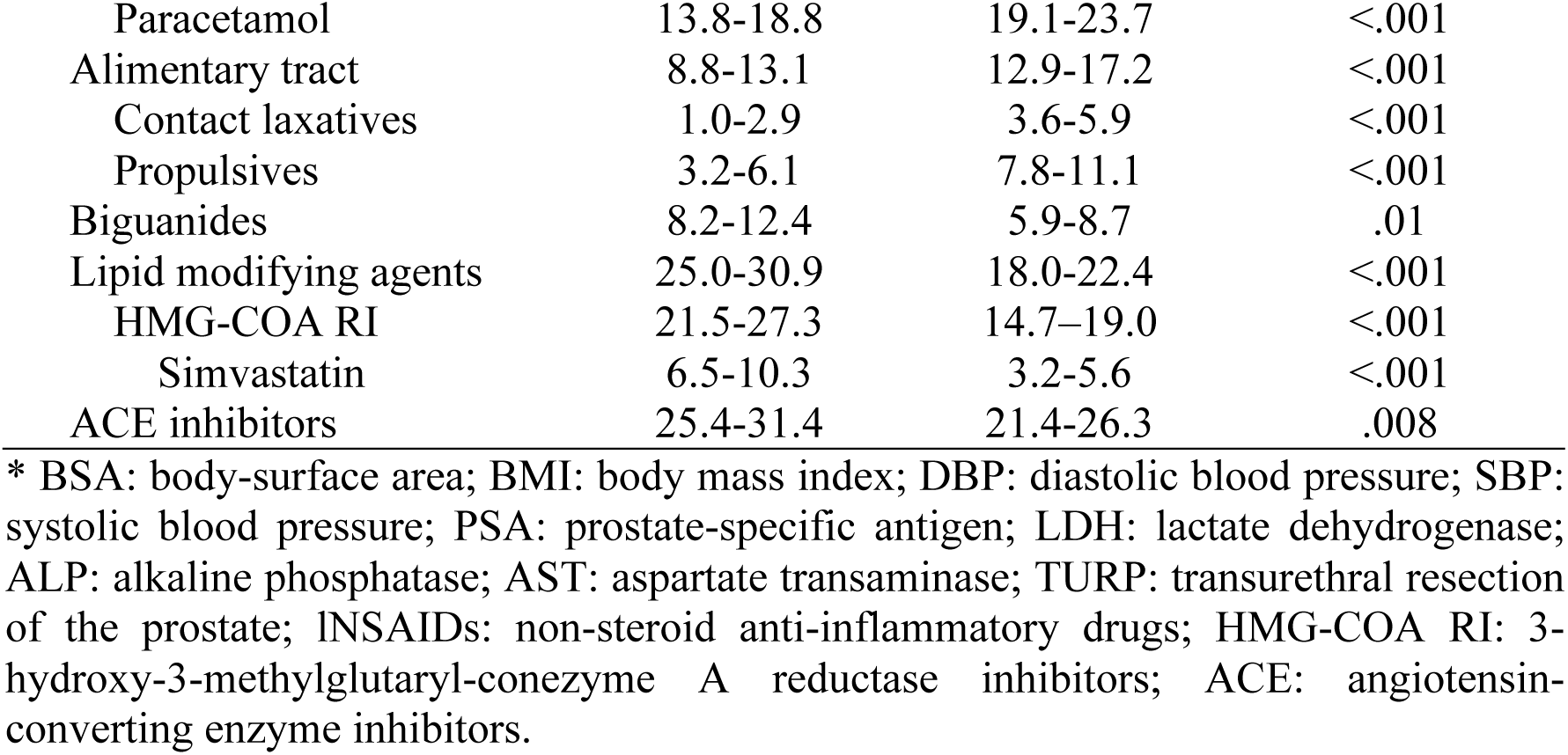
Significant differences in clinical and laboratory characteristics between subpopulations A (SPA) and B (SPB) as detected by an unsupervised machine learning model. All values correspond to confidence interval 95% (CI95%).

| Variable | SPA | SPB | p-value |
| --- | --- | --- | --- |
| N | 828 | 1,207 | - |
| N (%) | 40.6 | 59.4 | - |
| BSA* (m<sup>2</sup>) | 2.03-2.06 | 2.01-2.03 | .003 |
| BMI (Kg/m<sup>2</sup>) | 28.0-28.7 | 27.4-28.0 | .009 |
| Heart rate (bpm) | 71.6-74.9 | 74.8-77.5 | .007 |
| DBP (mmHg) | 76.2-77.8 | 78.4-80.3 | .001 |
| SBP (mmHg) | 130.8-133.5 | 133.5-136.6 | .006 |
| PSA (ng/mL) | 32.9-36.5 | 316.0-426.7 | <.001 |
| LDH (UI/L) | 186.0-192.0 | 291.0-328.0 | <.001 |
| Albumin (g/L) | 4.24-4.32 | 4.14-4.20 | <.001 |
| ALP (UI/L) | 92.5-97.4 | 330.3-378.1 | <.001 |
| AST (IU/L) | 23.6-24.8 | 29.3-31.2 | <.001 |
| Haemoglobin (g/L) | 12.8-13.0 | 12.1-12.3 | <.001 |
| ECOG |  |  |  |
| 0 (yes;%) | 54.1-60.6 | 39.3-44.8 | <.001 |
| 1 (yes;%) | 37.6-44.1 | 50.7-56.3 | <.001 |
| 2 (yes;%) | 1.10-3.00 | 3.20-5.70 | .001 |
| Bone-related signs and symptoms | 1.67-3.86 | 4.48-7.10 | <.001 |
| Bone pain | 3.07-5.85 | 7.33-10.5 | .008 |
| Therapy since diagnosis |  |  |  |
| Biphosphonates (total) | 17.0-22.4 | 22.5-27.4 | .005 |
| Zoledronic acid | 10.0-14.4 | 19.9-22.4 | <.001 |
| Opioids (total) | 23.0-28.9 | 39.5-45.1 | <.001 |
| Natural opium alkaloids | 7.80-11.8 | 15.3-19.6 | <.001 |
| Other opioids | 2.90-5.60 | 8.90-12.3 | <.001 |
| Opioids with other analgesics | 3.40-6.30 | 6.60-9.60 | .002 |
| Previous surgeries |  |  |  |
| Orchidectomy | 1.19-4.19 | 4.91-8.29 | <.001 |
| TURP | 3.46-7.34 | 0.07-0.89 | <.001 |
| Previous cancer therapies |  |  |  |
| Flutamide | 10.4-14.9 | 14.5-18.6 | .01 |
| Leuprolide | 36.4-43.1 | 30.7-36.0 | .003 |
| Opioids |  |  |  |
| Phenylpiperidine derivatives | 0.80-2.50 | 3.40-5.70 | <.001 |
| Fentanyl | 0.70-2.40 | 3.40-5.70 | <.001 |
| Morphine | 2.10-4.40 | 4.40-7.00 | .007 |
| Oxycodone | 3.30-6.10 | 7.40-10.6 | <.001 |
| Tramadol | 2.50-5.00 | 6.40-9.50 | <.001 |
| Concomitant medication |  |  |  |
| NSAIDs | 30.8-37.0 | 40.8-46.4 | <.001 |
| Diclofenac | 1.6-3.8 | 3.7-6.1 | .01 |
| Ibuprofen | 2.6-5.2 | 5.2-8.1 | .02 |
| Paracetamol | 13.8-18.8 | 19.1-23.7 | <.001 |
| Alimentary tract | 8.8-13.1 | 12.9-17.2 | <.001 |
| Contact laxatives | 1.0-2.9 | 3.6-5.9 | <.001 |
| Propulsives | 3.2-6.1 | 7.8-11.1 | <.001 |
| Biguanides | 8.2-12.4 | 5.9-8.7 | .01 |
| Lipid modifying agents | 25.0-30.9 | 18.0-22.4 | <.001 |
| HMG-COA RI | 21.5-27.3 | 14.7-19.0 | <.001 |
| Simvastatin | 6.5-10.3 | 3.2-5.6 | <.001 |
| ACE inhibitors | 25.4-31.4 | 21.4-26.3 | .008 |

\* BSA: body-surface area; BMI: body mass index; DBP: diastolic blood pressure; SBP: systolic blood pressure; PSA: prostate-specific antigen; LDH: lactate dehydrogenase; ALP: alkaline phosphatase; AST: aspartate transaminase; TURP: transurethral resection of the prostate; NSAIDs: non-steroid anti-inflammatory drugs; HMG-COA RI: 3-hydroxy-3-methylglutaryl-coenzyme A reductase inhibitors; ACE: angiotensin-converting enzyme inhibitors.

**Supplementary Table 5.** PSA, ALP and AST values for patients on the training set (TRS) and different validation datasets (VSs) included in the PROGRESS analysis.

| Dataset | Clinical Trial | N | Mean | CI95% |
| --- | --- | --- | --- | --- |
| TRS | NCT00273338, | 1,979 |  |  |
| PSA (ng/mL) | NCT00988208, |  | 237.2 | 203.2-271.1 |
| ALP (IU/L) | NCT00617669, |  | 243.5 | 258.2 |
| AST (IU/L) | NCT00519285 |  | 27.6 | 228.8-258.2 |
| VS1 | NCT00554229, | 531 |  |  |
| PSA (ng/mL) | NCT00676650 |  | 270.4 | 200.2-340.8 |
| ALP (IU/L) |  |  | 224.3 | 183.6-265.1 |
| AST (IU/L) |  |  | 27.0 | 25.8-29.2 |
| VS2 | TAX327, | 708 |  |  |
| PSA (ng/mL) | NCT00417079 |  | 392.3 | 303.7-481.0 |
| ALP (IU/L) |  |  | 339.6 | 280.7-398.6 |
| AST (IU/L) |  |  | 26.8 | 25.4-28.1 |
| VS3 | NCT00626548 | 660 |  |  |
| PSA (ng/mL) |  |  | 24.5 | 20.6-28.4 |
| ALP (IU/L) |  |  | 97.2 | 92.6-101.7 |
| AST (IU/L) |  |  | 24.7 | 23.9-25.4 |

\* PSA: prostate-specific antigen; ALP: alkaline phosphatase; AST: aspartate transaminase.

## SUPPLEMENTARY FIGURES

**Supplementary Figure 1.**
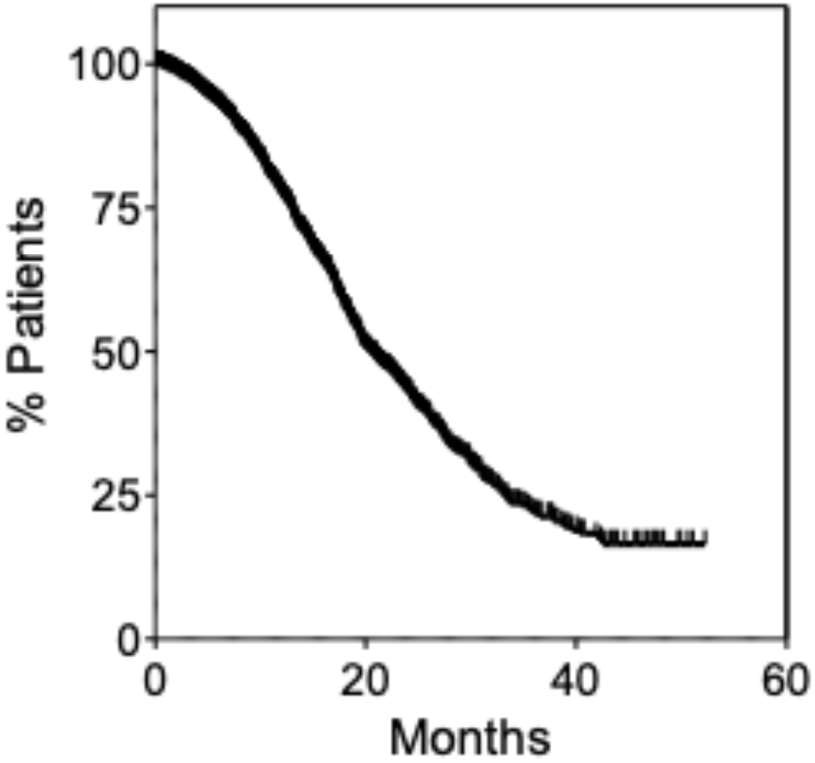
Kaplan-Meier estimates of OS of patients on the training set before clustering by an unsupervised machine learning model. Median OS for the total population (N=2,035) was 20.4 months (95%CI 19.4-21.8).

**Supplementary Figure 2.**
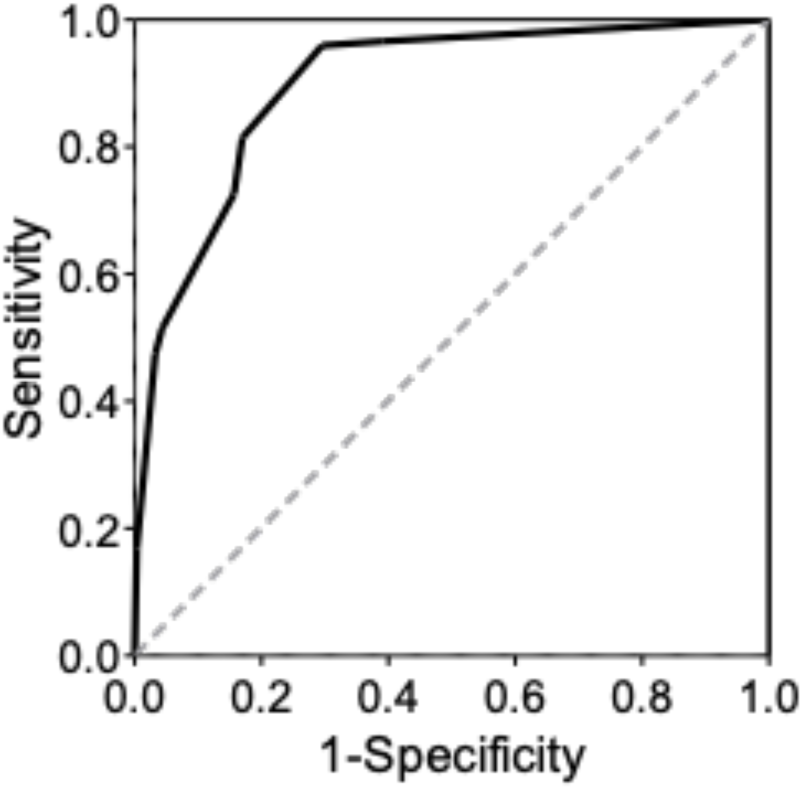
Receiver-Operating Characteristic Curve (ROC) of PROGRESS scoring system for predicting survival outcome.

**Supplementary Figure 3.**
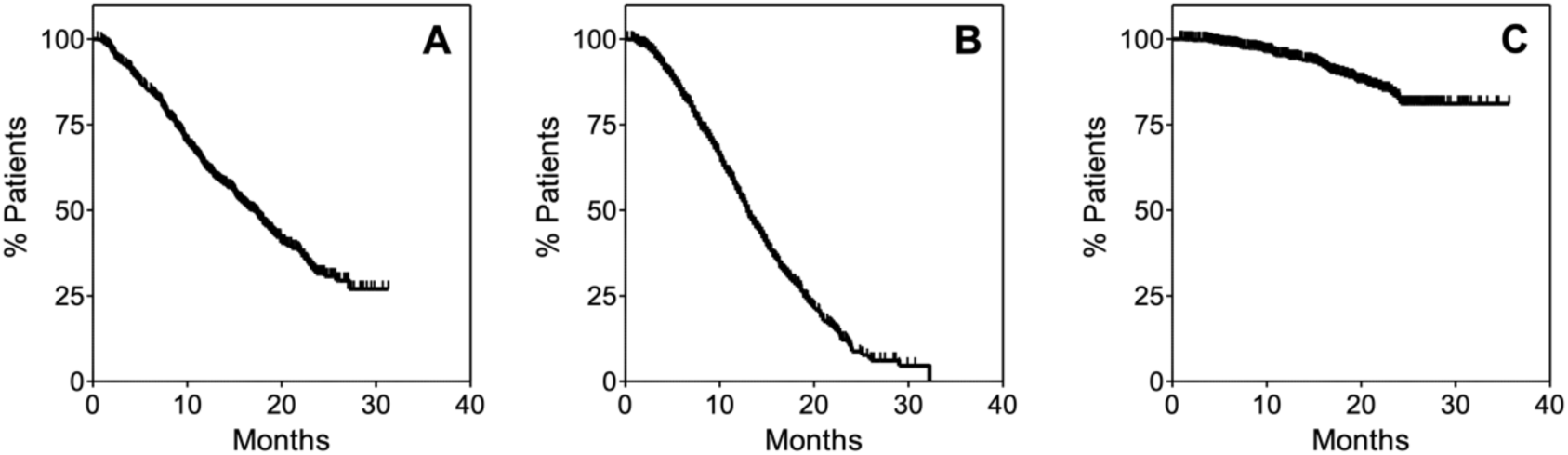
Kaplan-Meier estimates of OS of patients of the validation datasets. Panel (A) shows OS for VS1; median OS for the total population (N=531) was 16.9 months (95%CI: 15.1-18.4). Panel (B) shows OS for VS2; median OS for the total population (N=708) was 12.9 months (95%CI: 12.2-13.6). Panel (C) shows OS for VS3; Median OS was not reached during follow-up.

## Notes

### Funding Statement

This study did not receive any funding

### Author Declarations

The study used ONLY openly available human data that were originally located at: http://www.projectdatasphere.com/

